# Prevalence of mild behavioral impairment among a Chinese cohort of elderly with mild cognitive impairment and subjective cognitive impairment

**DOI:** 10.1101/2023.06.10.23291238

**Authors:** Yining Pan, You-qiang Song, Leung wing Chu, Yat-fung Shea

**Affiliations:** School of Biomedical Sciences, LKS Faculty of Medicine, The University of Hong Kong, Hong Kong, China; Department of Medicine, LKS Faculty of Medicine, The University of Hong Kong, Queen Mary Hospital, Hong Kong, China

## Abstract

**Introduction:** Mild behavioural impairment (MBI) describes later-life onset of sustained and meaningful neuropsychiatric symptoms (NPS) of any degree of severity before the onset of dementia or even the onset of cognitive symptoms. To our knowledge, there was only one study conducted in China and it only included non-dementia patients but did not differentiate among the cognitive groups.

**Methods:** A total of 88 subjects (28 cognitively normal, 15 subjective cognitive impairment, and 45 mild cognitive impairment) were recruited from Memory Clinic from 1st August 2020 through 16th Dec 2021. Inclusion criteria were that subjects should be of Chinese Han ethnicity. MBI was diagnosed according to the International Society to Advance Alzheimer’s Research and Treatment - Alzheimer’s Association (ISTAART-AA) criteria. MBI-checklist (MBI-C) score was either self-rated or informant-rated. NPI severity of 0 was defined as having a score of 0 for all Neuropsychiatric Inventory Questionnaire (NPI-Q) questions. NPI severity of 1 was given when the subject had a maximum score of 1-3. NPI severity of 2 referred to a subject having a score of four or over for at least one question. This cross-sectional study aimed to 1) calculate the prevalence of MBI among SCI/CN and MCI subjects in the Chinese population; 2) compare the performance of MBI-C and NPI-Q scales in MBI case ascertainment.

**Results:** Our results showed that the prevalence of MBI was 7% in CN/SCI group and 11.1% in MCI group assessed by MBI-C. The prevalence of NPS (severity >0) was 69% in CN/SCI group and 72.7% in MCI group assessed by NPI-Q. Spearman’s correlation test showed that the correlations of MBI and NPI severity were significant in overall subjects (correlation coefficient=0.3460, p=0.0011) and MCI group (correlation coefficient=0.3710, p=0.0131), and showing a trend of significance in CN/SCI group (correlation coefficient=0.3040, p=0.0503). However, Cohen’s kappa test showed that MBI-C and NPI-Q scale had only slight agreement in the overall and the both cognitive groups (overall: κ=0.0101; CN/SCI: κ=0.0667; MCI: κ=0.0917).

**Conclusion:** In conclusion, prevalence of MBI is around 7-11% among CN/SCI and MCI subjects in the Chinese population. MBI-C and NPI-Q were different tools and the former should be relied on to detect MBI. Further studies should validate an optimal cut-off score on MBI-C for each cognitive group in the Chinese population.

---

Mild behavioural impairment (MBI) describes later-life onset of sustained and meaningful neuropsychiatric symptoms (NPS) of any degree of severity before the onset of dementia or even the onset of cognitive symptoms.(1) Our previous systematic review and meta-analysis showed MBI is common among cognitively normal (CN), subjective cognitive impairment (SCI), and mild cognitive impairment (MCI) subjects, with a prevalence of 17%, 35.8%, and 45.5%, respectively.(2) MBI represents higher risk of cognitive deterioration.(2) To our knowledge, there was only one study conducted in China and it only included non-dementia patients but did not differentiate among the cognitive groups.(3)

Formal diagnostic criteria of MBI and a rating scale, MBI checklist (MBI-C) have been developed in 2017.(1) Since the MBI-C is relatively new and it is still undergoing validation among different cohorts, other tools such as Neuropsychiatric Inventory Questionnaire (NPI-Q) is being used to approximate MBI.(4) However, NPI-Q is used for assessing dementia population, and it requires a reference time of one month duration of symptoms, which is shorter than the six months requirement in MBI criteria. Therefore, there is a need to examine the agreement between MBI-C and NPI-Q on capturing MBI symptoms among our local population. This cross-sectional study aimed to 1) calculate the prevalence of MBI among SCI/CN and MCI subjects in the Chinese population; 2) compare the performance of MBI-C and NPI-Q scales in MBI case ascertainment.

A total of 88 subjects (28 CN, 15 SCI, and 45 MCI) were recruited from Memory Clinic from 1^st^ August 2020 through 16^th^ Dec 2021. Inclusion criteria were that subjects should be of Chinese Han ethnicity. MBI was diagnosed according to the International Society to Advance Alzheimer’s Research and Treatment - Alzheimer’s Association (ISTAART-AA) criteria.(1) There are no validated cut-offs for the MBI-C. Based on literature review, we used a total score of 8.5, 8.5, 6.5 as the cut-off for CN, SCI and MCI subjects, respectively.(5-7) MBI-C score was either self-rated or informant-rated. NPI severity of 0 was defined as having a score of 0 for all NPI questions. NPI severity of 1 was given when the subject had a maximum score of 1-3. NPI severity of 2 referred to a subject having a score of four or over for at least one question. MCI was diagnosed according to recommendations from the National Institute on Aging-Alzheimer’s Association workgroups in 2013. SCI was diagnosed by reporting a memory decline or cognitive function decline with worry (concern), in the context of normal HK-Montreal Cognitive Assessment Hong Kong Version score were considered having SCI. CN subjects did not meet either of the diagnostic criteria. Patients with major neurological disorders or other illness that could affect subjects’ scores on cognitive assessments were excluded. All statistical analysis were conducted with R version 4.0.4. A *p*-value less than 0.05 was considered statistically significant.

Our results showed that the prevalence of MBI was 7% in CN/SCI group and 11.1% in MCI group assessed by MBI-C. (Table 1) The prevalence of NPS (severity >0) was 69% in CN/SCI group and 72.7% in MCI group assessed by NPI-Q. Spearman’s correlation test showed that the correlations of MBI and NPI severity were significant in overall subjects (correlation coefficient=0.3460, *p*=0.0011) and MCI group (correlation coefficient=0.3710, *p*=0.0131), and showing a trend of significance in CN/SCI group (correlation coefficient=0.3040, *p*=0.0503). In the overall and both of the cognitive groups, MBI-C score and NPI-Q score were significantly correlated with each other (overall: correlation coefficient=0.8082, *p*<0.0001; CN/SCI: correlation coefficient=0.7337, *p*<0.0001; MCI: correlation coefficient=0.7715, *p*<0.0001). However, Cohen’s kappa test showed that MBI-C and NPI-Q scale had only slight agreement in the overall and the both cognitive groups (overall: κ=0.0101; CN/SCI: κ=0.0667; MCI: κ=0.0917).

**Table 1.** Characteristics of CN/SCI and MCI subjects.

|  | **CN/SCI**<br>**(n=43)** | **MCI (n=45)** | ***p*-value** |
| --- | --- | --- | --- |
| Age, yr | 72.2±8.9 | 77.5±8.3 | 0.0051* |
| Female sex, n (%) | 20 (46.5%) | 28 (62.2%) | 0.1390 |
| HK-MoCA |  |  |  |
| MBI (Yes), n (%) | 3 (7%) | 5 (11.1%) | 0.7139 |
| MBI, score | 2.9±5.3 | 3.2±6.1 | 0.7884 |
| NPI, score | 3.6±5.2 | 5.4±8.4 | 0.3452 |
| NPI, n (severity 0/1/2) <sup>§</sup> | 13/21/8 | 12/19/13 | 0.7325 |

<sup>§</sup>NPI-Q score was missing in one patient.
MBI: Mild Behavioural Impairment; NPI: Neuropsychiatric Inventory; HK-MoCA: Montreal Cognitive Assessment Hong Kong Version

Our results showed that MBI-C and NPI-Q were different in capturing MBI symptoms and the former tool should be used. The insignificant difference in the prevalence of MBI among both cognitive groups may because the cut-offs we employed were not specific enough to differentiate the prevalence of MBI among different cognitive groups. Further studies should be conducted to validate a cut-off of MBI-C in each cognitive group in the Chinese population. Our limited sample size and participants from one single centre were limitations of this study The prevalence of MBI is expected to be higher in a clinical setting than in the community, and thus our results may not be generalizable to other settings. In addition, due to the limited number of CN and SCI subjects, we had to combine the two groups together in the statistical analysis.

In conclusion, prevalence of MBI is around 7-11% among CN/SCI and MCI subjects in the Chinese population. MBI-C and NPI-Q were different tools and the former should be relied on to detect MBI. Further studies should validate an optimal cut-off score on MBI-C for each cognitive group in the Chinese population.

## Data Availability

All data produced in the present work are contained in the manuscript

## Authors’ declaration

Authors declared no conflict of interest.

## ACKNOWLEDGEMENTS

None.

